# Personal bias in nutrition advice: A survey of healthcare professionals’ recommendations regarding dairy and plant-based dairy alternatives

**DOI:** 10.1101/2021.07.20.21260856

**Authors:** Bridget E. Clark, Lizzy Pope, Emily H. Belarmino

## Abstract

**Objective:** To examine the relationship between health professionals’ personal dietary preferences and their professional nutrition advice on dairy and plant-based dairy alternatives.

**Design:** Cross sectional web-based survey. Survey examined health professionals’ personal dietary preferences, including milk preference, (plant-based or dairy milk), dietary pattern (vegetarian or omnivore), and if they followed a “plant-based” diet, and examined if they would advise patients to consume dairy and/or plant-based dairy alternatives. Logistic regression models examined if health professionals’ personal dietary preferences were associated with their willingness to recommend either product.

**Setting:** Sample of U.S. health professionals.

**Participants:** Non-probability convenience sampling recruited registered dietitian nutritionists, physicians, physician assistants, nurse practitioners, registered nurses, licensed practical nurses, dentists, dental hygienists, and students in any of these degree programs holding Junior standing and above. 331 respondents completed relevant survey questions and were included in analyses.

**Results:** Most respondents would recommend dairy (81%) and dairy alternatives (72%) to their patients. Half (49%) reported a preference for plant-based milk, about 40% identified as having a dietary pattern that reduces animal product intake, and about 40% reported following a plant-based dietary pattern. Plant-based milk preference (OR 4.52; 95% CI 2.31 to 8.82, *p*<0.001) and following a vegetarian dietary pattern (OR 1.91; 95% CI 1.11 to 3.27, *p*=0.019) were associated with greater odds of recommending dairy alternatives to a patient. Plant-based milk preference (OR 0.16; 95% CI 0.07 to 0.35, *p*<0.001), following a vegetarian dietary pattern (OR 0.45; 95% CI 0.25 to 0.82, *p*=0.009), and following a plant-based dietary pattern (OR 0.41; 95% CI 0.22 to 0.76, *p*=0.005) were associated with lessor odds of recommending dairy to a patient. Dietetics professionals were more likely than all other health professionals to recommend both dairy and dairy alternatives to patients.

**Conclusions:** Healthcare professionals’ nutrition recommendations appear to reflect their personal nutrition habits, despite dairy being an important source of essential nutrients for patients who are willing and able to consume it. Improved nutrition training focusing on evidence-based nutrition recommendations, reducing personal bias in practice, and routinely including registered dietitians on interprofessional healthcare teams may improve the quality of nutrition advice given to U.S. consumers.

## INTRODUCTION

Americans consider health professionals among the most trustworthy sources of health and nutrition information(1,2). Healthcare professionals are often called upon to advise patients on dietary choices, and are expected to provide sound nutrition recommendations using the most up to date evidence(3,4). Unfortunately, many health degree programs other than dietetics programs lack adequate nutrition education(3,5–7), making it difficult for health providers outside of registered dietitian nutritionists (RDNs) to accurately and confidently provide nutrition advice(6). Prior research also shows that health professionals’ personal health behaviors may influence the health advice they give to patients(8–12), and evidence suggests they are more confident in counseling patients on health and nutrition behaviors they practice themselves(13). Work examining this relationship has found that physical activity habits among health professionals and health professional degree students are likely to influence how much physical activity advice they provide(9,11,12). Likewise, in a study of U.S. female physicians from the Woman Physicians’ Health Study, those who were more concerned with their own eating habits or identified as a vegetarian were more likely to advise patients on nutrition(10). A study of Canadian physicians found those who consumed more fruits and vegetables advised patients on nutrition more often(9).

Health professionals may be asked by patients for advice on diets that consist mainly of plant-based (PB) foods. In recent years, many Americans have reported reducing their intake of animal-source foods(14,15). National and international reports have begun to highlight evidence showing the potential long-term health benefits of diets that emphasize vegetables, fruits, and whole grains, and limit intake of animal-source foods(16,17). Growing evidence demonstrates that PB dietary patterns are protective against multiple non-communicable diseases, including cardiovascular disease(18–20), type 2 diabetes(18,19), obesity(18), and certain cancers(18,19).

As Americans are reducing their animal product intake, sales of substitute products for popular animal-based foods has increased(21). Among the most popular are dairy alternatives, including PB milk, cheese, and yogurt products(21). PB dairy alternatives can be useful for individuals who have an allergy or intolerance to dairy(22), or who choose not to consume dairy for ethical reasons such as concerns for the environment or animal welfare(14). Many consumers also choose dairy alternatives due to perceived health benefits(14,23). However, evidence of health benefits from omitting dairy from the diet is inconsistent(24). Some research shows that dairy can have positive health impacts, potentially reducing one’s risk of type 2 diabetes and certain cancers(24). A diet of predominately plant foods may be protective against some chronic diseases(18), but the complete exclusion of a certain food groups may not be, and may lead to risk of nutrient inadequacies(19,20). Because PB dairy alternatives do not have equal nutrient content to dairy, especially in terms of protein and micronutrients(23,25,26), consumers who choose these products must ensure they are not missing these key nutrients in their diet.

It is important to assess how healthcare professionals advise patients on dairy and PB dairy alternatives, and if they provide unbiased recommendations based on the evidence of health outcomes associated with intake of these products. Accurate nutrition guidance from healthcare providers can positively impact patient dietary behavior(27). No research to date has examined if a relationship exists between healthcare professionals’ personal dietary preferences related to PB diets and their recommendations on dairy and dairy alternative products. The objectives of this study were to examine health professionals’ dietary preferences, which milk products (dairy and/or PB) they would recommend to patients and why, and if their personal preferences are associated with their professional advice on either product. We hypothesized that health professionals who follow animal product-reducing or PB dietary patterns or prefer PB milk to dairy milk would be more likely to recommend PB dairy alternatives and less likely to recommend dairy products to patients.

## METHODS

### SURVEY

We designed a web-based (Qualtrics, Provo, UT) survey to measure health professionals’ personal dietary preferences, and their recommendations on dairy products and PB dairy alternatives. To be eligible, individuals had to be age 18 years or older; live in the U.S. since at least January 2020; and currently be a registered dietitian nutritionist (RDN), medical doctor (MD), doctor of osteopathy (DO), physician assistant (PA), dentist, dental hygienist, licensed practical nurse (LPN), registered nurse (RN), nurse practitioner (NP), or a student currently enrolled in a degree program for one of those professions holding Junior undergraduate standing or above. These specific categories of health professionals were targeted as likely to have the most direct contact and opportunity to provide nutrition advice to patients. Our survey instrument was designed with input from nutrition and public health researchers with different professional expertise and dietary habits.

The survey asked respondents’ age, gender identity, race, ethnicity, state of residence, and type of health profession or professional degree program. To measure health professionals’ recommendations on dairy and PB alternatives, we included a 4-item instrument modified from two previous studies(28,29), which asked if respondents would recommend dairy foods to a patient and if they would recommend PB dairy alternatives to a patient. Response options for both questions were yes, no, and unsure. If respondents answered affirmatively to either question, they were asked which purpose(s) would lead them to recommend that product: maintenance of good health; prevention of chronic disease; treatment of chronic disease; nutrient deficiency; intolerance or allergy; or other with a space to elaborate.

We used a series of questions to measure health professionals’ dietary patterns. First, an instrument adapted from the National Health and Nutrition Examination Survey (NHANES) Dietary Screener Questionnaire (DSQ)(30,31) asked respondents the major type of milk they consume, and the frequency they consume it. Five response options were provided: from cows, from goats or sheep, PB milk, other, or I don’t drink milk. Those who responded other could write in the type of milk consumed. We then asked which type of dietary pattern respondents most closely identify with: vegan (not consuming any animal products); vegetarian (not consuming any meat or fish); semi-vegetarian (consuming red meat, poultry or fish no more than once a week); pesco-vegetarian (consuming no meat but fish); omnivorous (eating meat or fish almost every day); or other with a space to write-in their dietary pattern(32). Lastly, we asked respondents whether they consider their diet to be “plant-based” to which they could respond affirmatively or negatively.

To reduce bias that could be introduced through utilizing a single method of non-probability convenience sampling, we used three methods to recruit respondents: (1) paid digital ads via Facebook to reach a national sample; (2) targeted postings to relevant national LinkedIn and Facebook group pages; and (3) postings on academic, professional, and community listservs in Vermont as well as nationally. The survey was open from November 19^th^, 2020 until February 4, 2021 and 417 people completed at least part of the survey. For this analysis, we excluded 86 respondents who did not answer both recommendation questions, resulting in a final sample of 331. Respondents were entered in a gift card drawing. This study was determined to be exempt by the Institutional Review Board at the University of Vermont.

### ANALYSIS

We analyzed survey data with IBM SPSS Statistics (version 27). During analysis, variables for race and ethnicity were merged and recoded to a binary variable “Black, Indigenous, or person of color (BIPOC)” and “not BIPOC”. State of residence was recoded to a binary variable that indicated whether it was a “dairy state”. U.S. dairy states were determined based on being among the 10 states with the highest percentage of total farm sales coming from milk sales in 2017 (VT, NM, NY, WI, ID, NH, PA, AZ, MI, and ME)(33). For milk product preference, respondents who reported other and could not be reclassified or reported not drinking milk were excluded from analyses that included this variable, as an aim of this study was to examine preference for dairy versus PB alternative products. Dietary pattern was recoded to a binary omnivorous/vegetarian variable, with vegetarians representing all respondents who reported a dietary pattern that reduced meat intake. Respondents who selected other as their dietary pattern but described a diet that fit into one of the specified diets were reclassified to that category. Only two respondents could not be reclassified and were excluded from analyses that included this variable.

We generated univariate descriptive statistics for all variables. Using a series of six logistic regression models, we examined whether milk preference, dietary pattern, or considering one’s diet to be PB was associated with whether the respondent would recommend (1) dairy foods and (2) PB dairy substitutes. For the models, respondents who selected no or unsure regarding if they would recommend dairy and/or dairy alternatives were combined and compared to respondents who selected yes, they would recommend dairy and/or dairy alternatives. Following the construction of unadjusted models, we adjusted the models for age, whether or not the respondent lived in a dairy state, and health professional type (dietetics professional or student versus other type of health professional or student). Gender identity and race/ethnicity were excluded from the models due to the low number of participants who identified with a gender other than female or with a race other than non-Hispanic white. Health professional type was added to the models because a separate analysis with this dataset found perceptions of dairy and PB alternatives to differ between dietetics professionals/students and other types of healthcare professionals/students(34). We used listwise deletion to handle missing data. Tests were statistically significant if p < 0.05.

## RESULTS

Most survey respondents were female (91.5%), non-Hispanic white (87.5%), and under age 55 (82.2%) (Table 1). 46% were from a dairy state, reflecting the additional recruitment conducted in Vermont. The sample consisted of 276 practicing health professionals and 55 students hereafter collectively referred to as “health professionals”. RDNs and dietetics students made up 45.3% of the sample, and other health professionals or students made up the remainder. Nurses or nursing students comprised the largest population of other health professionals (71.8%; data not shown).

**Table 1:**
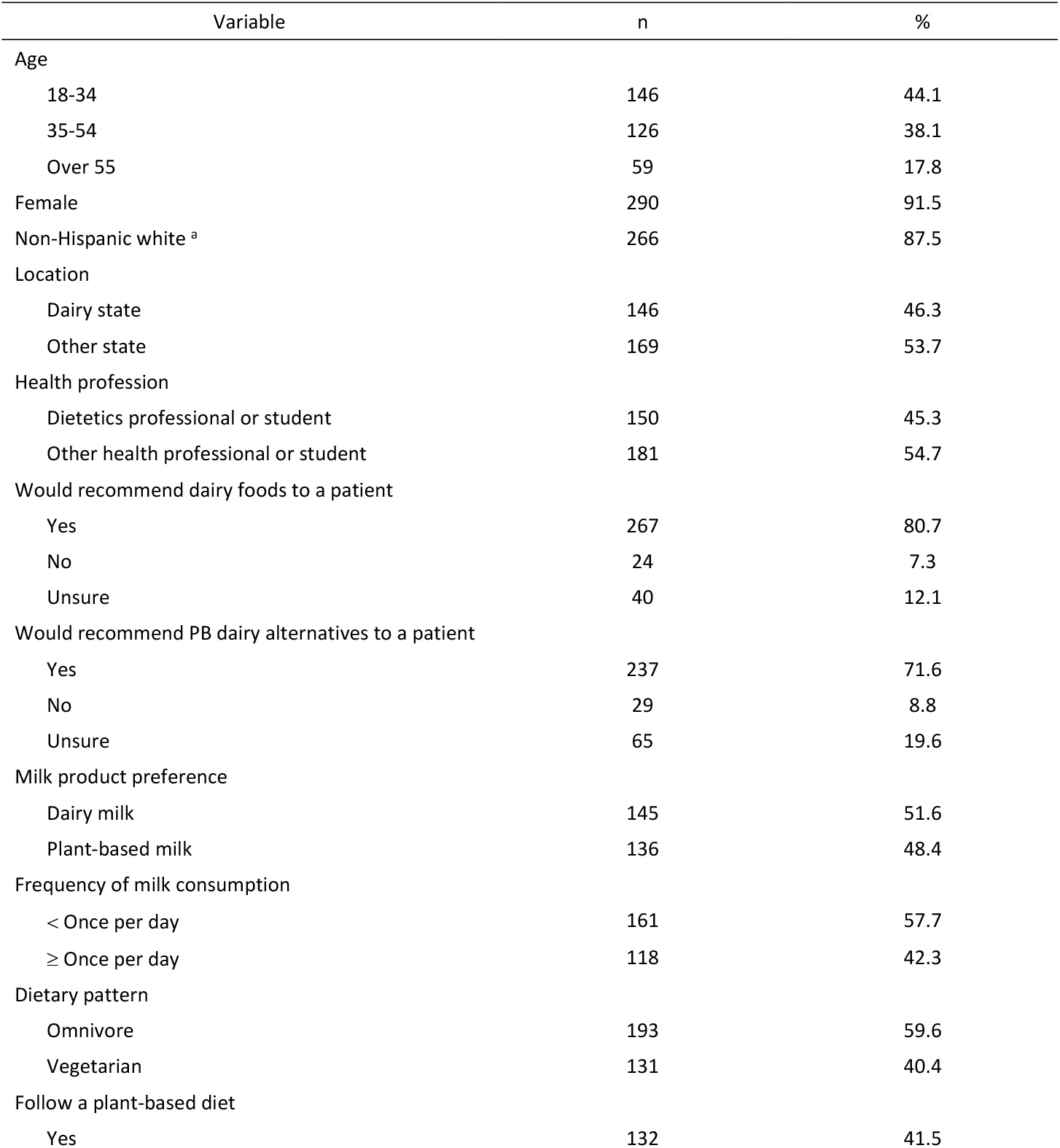
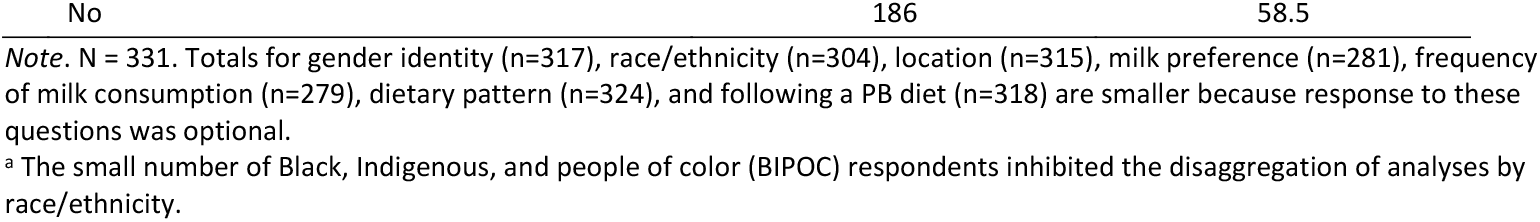
Characteristics of survey respondents.

### HEALTH PROFESSIONALS’ RECOMMENDATIONS ON DAIRY AND DAIRY ALTERNATIVES

The majority of health professionals would recommend both dairy (80.7%) and PB dairy alternatives (71.6%) to their patients (Table 1). Top reasons health professionals would recommend dairy foods to a patient included maintenance of good health (79.8%), a nutrient deficiency (73.7%), or a nut or soy allergy or intolerance (69.8%) (Figure 1). Dairy allergy or intolerance far outweighed all other reasons health professionals would recommend PB dairy alternatives to a patient, with almost all (95.3%) citing this reason (Figure 2). Of the 19.9% who stated “other” for why they would recommend dairy alternatives, almost half said the patient’s personal preference.

**Figure 1:**
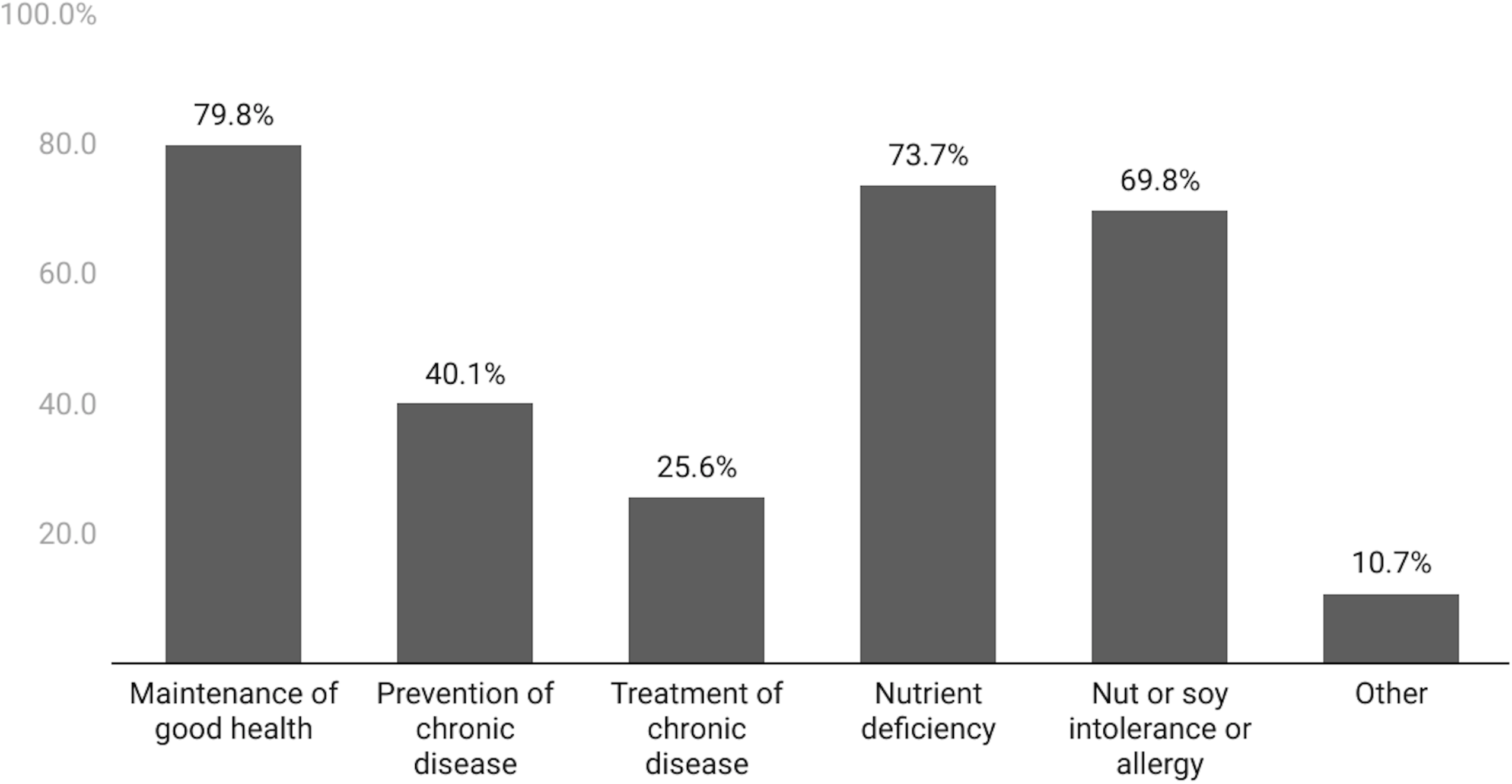
Percentage of respondents who would recommend dairy foods to a patient for each purpose. Percentages only include respondents who selected that they would recommend dairy foods to a patient (n=262).

**Figure 2:**
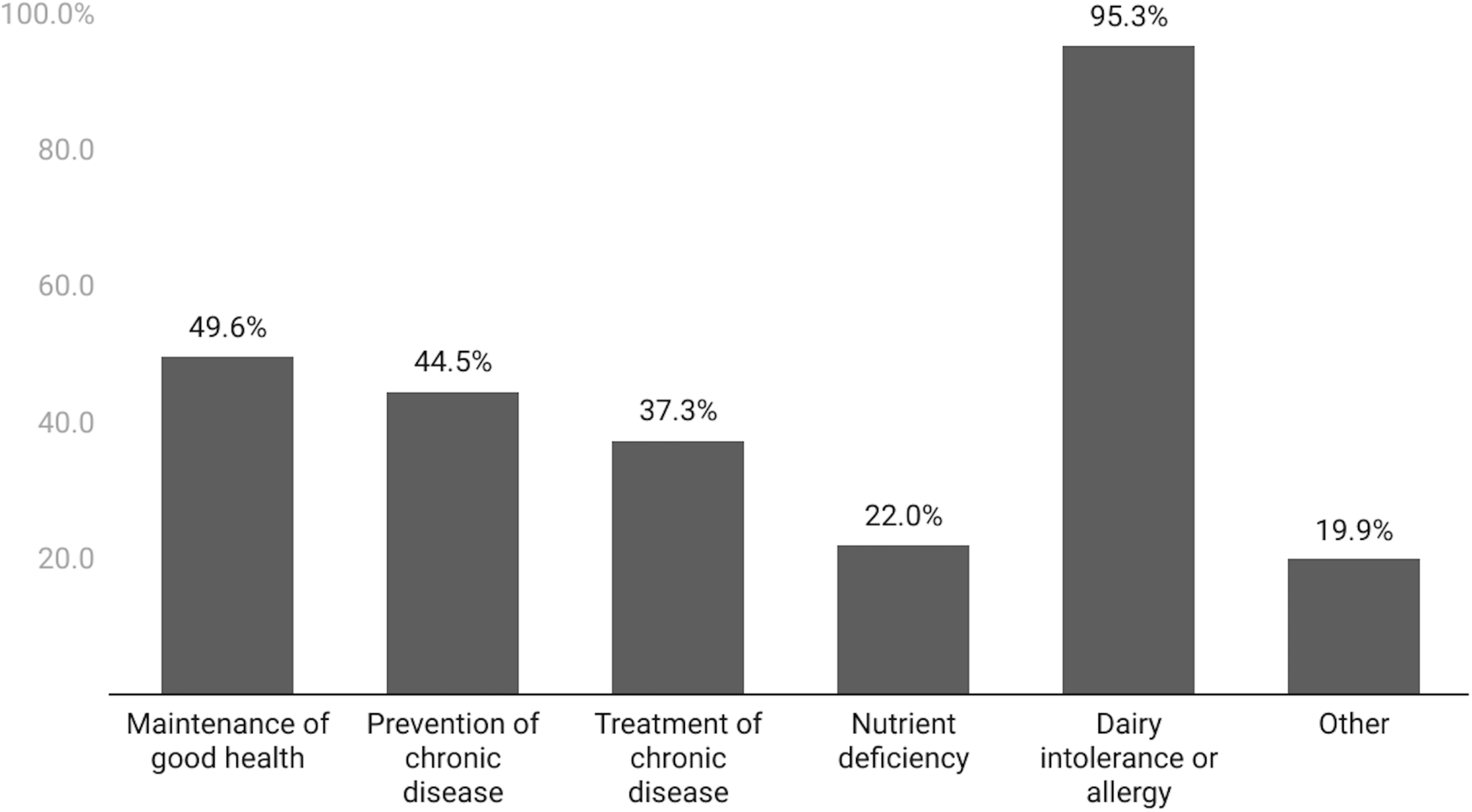
Percentage of respondents who would recommend PB dairy alternatives to a patient for each purpose. Percentages only include respondents who selected that they would recommend PB dairy alternatives to a patient (n=236).

### HEALTH PROFESSIONALS’ DIETARY PATTERNS

Almost half of respondents (48.4%) consume PB milk alternatives over dairy milk (Table 1). A little over half (57.7%) drink dairy or PB milk infrequently (less than once a day), and 42.3% drink milk frequently (once a day or more). About 60% of respondents reported following an omnivorous dietary pattern and about 40% reported following some form of a vegetarian diet. Likewise, about 40% of respondents considered their diet to be PB.

### HEALTH PROFESSIONALS’ MILK PRODUCT RECOMMENDATIONS BY PERSONAL DIETARY PATTERN

In unadjusted regression models, preference for PB milk (OR 0.14; 95% CI 0.06 to 0.30, *p*< 0.001) and identifying as vegetarian (OR 0.53; 95% CI 0.30 to 0.92, *p*=0.025) were significantly associated with lessor odds of recommending dairy foods to patients. Preference for PB milk (OR 3.76; 95% CI 2.09 to 6.79, *p*< 0.001), identifying as a vegetarian (OR 2.05; 95% CI 1.22 to 3.46, *p=*0.007), and following a PB diet (OR 1.90; 95% CI 1.14 to 3.19, *p*= 0.015) were significantly associated with greater odds of recommending dairy alternatives to patients (Table 2).

**Table 2:** Unadjusted regression models examining whether respondents would recommend dairy and PB dairy alternative products to their patients from personal preferences and dietary pattern.

| Model | Variable | Would recommend dairy products to a patient |  |  |  |  |
| --- | --- | --- | --- | --- | --- | --- |
|  |  | b | SE | OR | CI | p value |
| 1 | Milk preference (ref=milk from animals) |  |  |  |  |  |
|  | Plant-based milk | -1.98 | 0.39 | 0.14 | 0.06-0.30 | **< 0.001** |
| 2 | Dietary pattern (ref=omnivore) |  |  |  |  |  |
|  | Vegetarian | -0.64 | 0.29 | 0.53 | 0.30-0.92 | **0.025** |
| 3 | Follow a plant-based diet (ref=no) |  |  |  |  |  |
|  | Yes | -0.55 | 0.29 | 0.58 | 0.33-1.02 | 0.058 |
|  |  | Would recommend dairy alternatives to a patient |  |  |  |  |
|  |  | b | SE | OR | CI | p value |
| 4 | Milk preference (ref=milk from animals) |  |  |  |  |  |
|  | Plant-based milk | 1.33 | 0.30 | 3.76 | 2.09-6.79 | **< 0.001** |
| 5 | Dietary pattern (ref=omnivore) |  |  |  |  |  |
|  | Vegetarian | 0.72 | 0.27 | 2.05 | 1.22-3.46 | **0.007** |
| 6 | Follow a plant-based diet (ref=no) |  |  |  |  |  |
|  | Yes | 0.64 | 0.26 | 1.90 | 1.14-3.19 | **0.015** |

Note. The logistic regression models presented above used "no or unsure" as the reference category of the dependent variables. Milk preference n=281; Dietary pattern n=324; Follow a PB diet n=318.

In adjusted regression models, dietary preferences and health profession were associated with respondents’ willingness to recommend dairy and PB dairy alternatives to patients. In the first adjusted model examining milk preference (Table 3), health professionals who prefer PB milk were significantly less likely to say they would recommend dairy products to a patient (OR 0.16; 95% CI 0.07 to 0.35, *p*<0.001) and more likely to say they would recommend dairy alternatives to patient (OR 4.52; 95% CI 2.31 to 8.82, *p*<0.001). Frequent milk product drinkers were significantly less likely say they would recommend dairy alternatives to a patient (OR 0.47; 95% CI 0.26 to 0.86, *p*=0.014). As compared to non-dietetics professionals, dietetics professionals were significantly more likely to say they would recommend both dairy (OR 2.85; 95% CI 1.36 to 5.95, *p*=0.005) and dairy alternatives (OR 2.79; 95% CI 1.49 to 5.25, *p*=0.001) to a patient.

**Table 3:** Adjusted regression examining if respondents who prefer PB milk over dairy would recommend dairy and PB dairy alternative products to their patients (n=272).

| Variable | Would recommend dairy products to a patient |  |  |  |  | Would recommend dairy alternatives to a patient |  |  |  |  |
| --- | --- | --- | --- | --- | --- | --- | --- | --- | --- | --- |
|  | b | SE | OR | CI | p value | b | SE | OR | CI | p value |
| Age (ref=18-34) |  |  |  |  |  |  |  |  |  |  |
| 35-54 | -0.16 | 0.40 | 0.85 | 0.39-1.86 | 0.684 | 0.35 | 0.35 | 1.41 | 0.71-2.82 | 0.328 |
| Over 55 | -0.80 | 0.52 | 0.92 | 0.33-2.58 | 0.878 | 0.03 | 0.42 | 1.03 | 0.46-2.32 | 0.948 |
| Location (ref=Not Dairy State) |  |  |  |  |  |  |  |  |  |  |
| Dairy state | 0.33 | 0.36 | 1.39 | 0.68-2.81 | 0.365 | -0.24 | 0.30 | 0.79 | 0.44-1.43 | 0.436 |
| Health professional type (ref=non-dietetics professional) |  |  |  |  |  |  |  |  |  |  |
| Dietetics professional | 1.05 | 0.38 | 2.85 | 1.36-5.95 | **0.005** | 1.03 | 0.32 | 2.79 | 1.49-5.25 | **0.001** |
| Milk frequency (ref= < once per day) |  |  |  |  |  |  |  |  |  |  |
| ≥ once per day | -0.01 | 0.35 | 0.99 | 0.50-1.94 | 0.971 | -0.75 | 0.31 | 0.47 | 0.26-0.86 | **0.014** |
| Milk preference (ref=milk from animals) |  |  |  |  |  |  |  |  |  |  |
| Plant-based milk | -1.85 | 0.41 | 0.16 | 0.07-0.35 | **< 0.001** | 1.51 | 0.34 | 4.52 | 2.31-8.82 | **< 0.001** |

Note. The multivariate logistic regression models presented above used “no or unsure” as the reference category of the dependent variables.

The second adjusted model (Table 4), examining recommendations by dietary pattern, indicated that health professionals who identified as vegetarian were significantly less likely to say they would recommend dairy products to a patient (OR 0.45; 95% CI 0.25 to 0.82, *p*=0.009), and more likely to say they would recommend dairy alternatives to a patient (OR 1.91; 95% CI 1.11 to 3.27, *p*=0.019) as compared to health professionals who identified as omnivorous. Dietetics professionals were again significantly more likely to say they would recommend both dairy (OR 1.40; 95% CI 2.03 to 8.03, *p*<0.001) and dairy alternatives (OR 2.01; 95% CI 1.18 to 3.43, *p*=0.010) to a patient.

**Table 4:** Adjusted regression examining if respondents who follow a vegetarian diet would recommend dairy and PB dairy alternative products to their patients (n=314).

| Variable | Would recommend dairy products to a patient |  |  |  |  | Would recommend dairy alternatives to a patient |  |  |  |  |
| --- | --- | --- | --- | --- | --- | --- | --- | --- | --- | --- |
|  | b | SE | OR | CI | p value | b | SE | OR | CI | p value |
| Age (ref=18-34) |  |  |  |  |  |  |  |  |  |  |
| 35-54 | 0.26 | 0.34 | 1.30 | 0.66-2.55 | 0.447 | 0.08 | 0.30 | 1.09 | 0.60-1.96 | 0.784 |
| Over 55 | 0.45 | 0.44 | 1.57 | 0.66-3.75 | 0.311 | -0.38 | 0.35 | 0.68 | 0.34-1.36 | 0.276 |
| Location (ref=Not Dairy State) |  |  |  |  |  |  |  |  |  |  |
| Dairy state | 0.54 | 0.32 | 1.72 | 0.92-3.23 | 0.089 | -0.43 | 0.27 | 0.65 | 0.39-1.10 | 0.108 |
| Health professional type (ref=non-dietetics professional) |  |  |  |  |  |  |  |  |  |  |
| Dietetics professional | 1.40 | 0.35 | 4.03 | 2.03-8.03 | **< 0.001** | 0.70 | 0.27 | 2.01 | 1.18-3.43 | **0.010** |
| Dietary pattern (ref= Omnivore) |  |  |  |  |  |  |  |  |  |  |
| Vegetarian | -0.80 | 0.31 | 0.45 | 0.25-0.82 | **0.009** | 0.64 | 0.28 | 1.91 | 1.11-3.27 | **0.019** |

Note. The multivariate logistic regression models presented above used “no or unsure” as the reference category of the dependent variables.

**Table 4:** Adjusted regression examining if respondents who consider their diet to be PB would recommend dairy and PB dairy alternative products to their patients (n=309).

| Variable | Would you recommend dairy products to a patient |  |  |  |  | Would you recommend dairy alternatives to a patient |  |  |  |  |
| --- | --- | --- | --- | --- | --- | --- | --- | --- | --- | --- |
|  | b | SE | OR | CI | p value | b | SE | OR | CI | p value |
| Age (ref=18-34) |  |  |  |  |  |  |  |  |  |  |
| 35-54 | 0.17 | 0.35 | 1.19 | 0.60-2.36 | 0.618 | 0.14 | 0.30 | 1.15 | 0.63-2.07 | 0.653 |
| Over 55 | 0.43 | 0.44 | 1.54 | 0.64-3.67 | 0.334 | -0.42 | 0.35 | 0.66 | 0.33-1.32 | 0.240 |
| Location (ref=Not Dairy State) |  |  |  |  |  |  |  |  |  |  |
| Dairy state | 0.53 | 0.32 | 1.70 | .91-3.21 | .099 | -0.41 | 0.27 | 0.66 | 0.39-1.11 | 0.120 |
| Health professional type (ref=non-dietetics professional) |  |  |  |  |  |  |  |  |  |  |
| Dietetics professional | 1.54 | 0.36 | 4.65 | 2.29-9.45 | **< 0.001** | 0.60 | 0.28 | 1.82 | 1.06-3.12 | **0.031** |
| Follow a plant-based diet (ref= No) |  |  |  |  |  |  |  |  |  |  |
| Yes | -0.90 | 0.32 | 0.41 | 0.22-0.76 | **0.005** | 0.51 | 0.28 | 1.67 | 0.97-2.89 | 0.067 |

Note. The multivariate logistic regression models presented above used “no or unsure” as the reference category of the dependent variables.

The third adjusted model (Table 5) found respondents who reported that their diet was PB were significantly less likely to say they would recommend dairy products to a patient (OR 0.41; 95% CI 0.22 to 0.76, *p*=0.005). Dietetics professionals were significantly more likely to say they would recommend both dairy (OR 4.65; 95% CI 2.29 to 2.45, *p*<0.001) and dairy alternatives (OR 1.82; 95% CI 1.06 to 3.12, *p*=0.031) to a patient.

## DISCUSSION

To our knowledge this is the first paper to explore health professionals’ nutrition recommendations on dairy and PB dairy alternatives, and the first to examine if personal preferences for PB versus dairy milk or dietary pattern are associated with recommendations health professionals provide to consumers. As the consumption of PB dairy alternatives increases(21), it is important to understand if health professionals are providing accurate and unbiased nutrition advice about these products to their patients. Most health professionals sampled would recommend both dairy and dairy alternative products to patients, although their health reasons for recommending each product differed. Almost half of respondents preferred PB milk over dairy milk, and about 40% identified as vegetarian or described their diet as PB. Personal dietary preferences were associated with health professionals’ likelihood of recommending dairy and dairy alternatives. Health professionals who preferred PB milk or followed PB dietary patterns were more likely to recommend dairy alternatives and less likely to recommend dairy.

Maintenance of good health, nutrient deficiency, and nut or soy allergy or intolerance were major reasons health professionals would recommend dairy to a patient, while dairy allergy or intolerance far exceeded all other reasons for health professionals recommending PB dairy alternatives. Our findings agree with prior research indicating dairy allergy or intolerance is a top reason consumers choose PB alternatives(22). Aligning with current U.S. dietary recommendations(35), a much greater proportion would recommend dairy (compared to PB alternatives) for maintenance of good health. Few health professionals would recommend PB dairy alternatives to combat a nutrient deficiency and less than 50% said they would recommend dairy or a PB product for prevention and/or treatment of chronic disease. This may indicate that many health professionals don’t feel that milk products have a large influence on disease risk. Current literature(24) suggests dairy intake is neither the superfood nor the dietary villain that different stakeholders make it out to be(36,37).

The proportion of health professionals who indicated that they prefer to drink PB milk alternatives (48.4%) is higher than the estimated 23% of U.S. households that consume mostly non-dairy milks, as found in a recent representative study of 995 U.S. households(38). It is interesting that almost half prefer PB milk, as almost half of the sample came from a dairy state where dairy milk consumption may be expected to be higher. This high proportion is also interesting because most of the sample was made up of non-Hispanic white respondents, and this population is more likely to meet U.S. dairy intake recommendations(39). About 40% of health professionals identified as following a dietary pattern that reduces animal product intake to some extent, and about 40% consider their diets to be PB. Although only about 3-5% of Americans identify as vegetarian or vegan(40,41), over 40% report trying to consume more PB foods(39). Prior work has shown mixed results regarding whether the dietary behaviors of health professionals differ from the general public(7,42,43). Globally, less than 40% of physicians appear to consume more than two servings of fruits and vegetables per day, although over three-fourths appear to consume meat and dairy daily(7). Another study found that although U.S. female nurses do not consume the recommended amount of fruits and vegetables per day, their intake is higher than women in the general population(42). Further research may be required to determine the degree that health professionals and all Americans are fully or partially replacing animal-based foods with plant sources.

Health professionals who personally prefer PB milk or identify as vegetarian were more likely to say they would recommend PB dairy alternatives to patients, and less likely to say they would recommend dairy. Health professionals who consider their diet to be PB were also less likely to say they would recommend dairy. In unadjusted models, health professionals with PB diets were also more likely to recommend PB dairy alternatives, although when models were adjusted, this association was no longer significant. Our findings agree with the literature that personal nutrition behaviors are associated with health professionals’ nutrition recommendations(9,10). Although most respondents did report that they would recommend both dairy and PB dairy alternatives to patients, our findings suggest bias in nutrition advising. Whether dairy or a PB dairy alternative is appropriate for a given patient often depends on that patient’s individual dietary needs and preferences(14,22). While the associations between dairy intake and some health outcomes remain unclear(24), dairy is a top food source of many nutrients of concern(34), and advising a patient to choose a PB alternative in place of dairy if there is no personal or health-related need could lead to nutrition risks(19,20). The nutrition education provided in many health degree programs remains limited (3,5-7). Improved access to information regarding the nutritional characteristics of both dairy and PB alternatives may increase health professionals’ knowledge on this topic regardless of their own dietary practices. The greater propensity of RDNs to recommend either product as compared to other health professionals, suggests that their specialized training in nutrition and nutrition counseling may buffer some of the influence of personal preferences. Previous work has highlighted a need to increase interprofessional teamwork between different health professionals(3). Given the high rates of diet-related non-communicable diseases in the U.S.(44) and the high demand for nutrition information by consumers(45), RDNs represent critical members of interprofessional healthcare teams. Future work may focus on education models centered on collaboration between RDNs and other healthcare disciplines.

This study has several limitations. First, the relatively small sample size did not allow for certain statistical comparisons, such as the milk product recommendations between vegetarians, vegans, and other meat-reducing dietary patterns separately. However, this may be adequate, given that dietary pattern reporting is often imprecise(46). Second, few respondents reported that they would not recommend dairy or PB alternatives to their patient, hindering our ability to examine factors that associated with reluctance to recommend these products. Third, health professionals’ dietary patterns and preferences were based on self-reported data, which are subject to response bias(47). What was defined as a PB diet was also left to the respondents’ interpretation. Future research on this topic may consider more robust or objective measures of dietary patterns, such as dietary recalls or biomarkers. Fourth, the majority of the sample was female and non-Hispanic white, and nearly 85% were dietetics or nursing professionals. This reflects the additional recruitment activities targeted towards RDNs and nurses, and the extended recruitment in Vermont. Over 90% of RDNs and nurses in the U.S. are female(48,49) and less than 10% of the population of Vermont identify with a race or ethnicity other than non-Hispanic white(50). Most prior research examining associations between personal beliefs and behaviors and professional counseling practices has focused on physicians and medical students(8–10). This study was novel in that it examined counseling bias among other types of health professionals, and among those with and without formal dietetics training, which has not been examined previously.

## CONCLUSION

Our findings indicate that the personal milk preferences and dietary patterns of U.S. health professionals may be associated with willingness to recommend dairy and PB dairy alternatives to patients. Health professionals are expected to base their nutrition recommendations on up-to-date nutrition literature. While PB dairy alternative products are important substitutes for individuals who cannot or will not consume dairy, the needs of the patient should drive nutrition recommendations rather than health professionals’ personal beliefs. Future work should focus on strategies for improving nutrition training for healthcare professionals. Specifically, training could focus on use of evidence-based nutrition recommendations, reducing personal bias in nutrition advice, and the importance of referring patients with nutrition questions and concerns to RDNs and including RDNs on healthcare teams. This may help improve the quality of nutrition advice given to U.S. consumers.

## Data Availability

Data are available from the corresponding author on reasonable request.

## Data availability statement

Data are available from the corresponding author on reasonable request.

## Author contributions

All authors designed the study. BEC was responsible for analyzing the survey data. BEC and EHB contributed to the interpretation of the data. BEC drafted the manuscript. All authors contributed to manuscript revisions and read and approved the final manuscript.

## Patient consent for publication

Not required.

## Ethics approval

This study was approved by the Committee on Human Research in the Behavioral and Social Sciences (CHRBSS), the Institutional Review Board (IRB) of the University of Vermont.

## References

1. Funk C, Hefferon M, Kennedy B, et al. Trust and Mistrust in Americans’ Views of Scientific Experts. Pew Research Center, 2019. Available: https://www.pewresearch.org/science/2019/08/02/trust-and-mistrust-in-americans-views-of-scientific-experts/ [Accessed 4 Mar 2021].

2. Brenan M. Nurses Again Outpace Other Professions for Honesty, Ethics. Gallup News, 2018. Available: https://news.gallup.com/poll/245597/nurses-again-outpace-professions-honesty-ethics.aspx?g_source=link_NEWSV9&g_medium=NEWSFEED&g_campaign=item_&g_content=Nurses%20Again%20Outpace%20Other%20Professions%20for%20Honesty%2C%20Ethics [Accessed 4 Mar 2021].

3. DiMaria-Ghalili RA, Mirtallo JM, Tobin BW, et al. Challenges and opportunities for nutrition education and training in the health care professions: Intraprofessional and interprofessional call to action. Am J Clin Nutr 2014;99(Suppl 5):P1184S–93S.doi:10.3945/ajcn.113.073536

4. Anderson D, Baird S, Bates T, et al. Academy of nutrition and dietetics: Revised 2017 scope of practice for the registered dietitian nutritionist. J Acad Nutr Diet 2018;118(1):141– 65.doi:10.1016/j.jand.2017.10.002

5. Adams KM, Scott Butsch W, Kohlmeier M. The state of nutrition education at US medical schools. J Biomed Educ 2015;2015(4):1–7.doi:10.1155/2015/357627

6. Crowley J, Ball L, Hiddink GJ. Nutrition in medical education: A systematic review. Lancet Planet Health 2019;3(9):e379–89.doi:10.1016/S2542-5196(19)30171-8

7. Aggarwal M, Devries S, Freeman AM, et al. The deficit of nutrition education of physicians. Am J Med 2018;131(4):339–45.doi:10.1016/j.amjmed.2017.11.036

8. Frank E, Carrera JS, Elon L, et al. Predictors of US medical students’ prevention counseling practices. Prev Med (Baltim) 2007;44(1):76–81.doi:10.1016/j.ypmed.2006.07.018

9. Frank E, Segura C, Shen H, et al. Predictors of Canadian physicians’ prevention counseling practices. Can J Public Health 2010;101(5):390–5.doi:10.1007/bf03404859

10. Frank E, Wright EH, Serdula MK, et al. Personal and professional nutrition-related practices of US female physicians. Am J Clin Nutr 2002;75(2):326–32.doi:10.1093/ajcn/75.2.326

11. Lobelo F, Duperly J, Frank E. Physical activity habits of doctors and medical students influence their counselling practices. Br J Sports Med 2009;43(2):89–92.doi:10.1136/bjsm.2008.055426

12. Abramson S, Stein J, Schaufele M, et al. Personal exercise habits and counseling practices of primary care physicians: A national survey. Clin J Sport Med 2000;10(1):40–8. doi:10.1097/00042752-200001000-00008

13. Vickers KS, Kircher KJ, Smith MD, et al. Health behavior counseling in primary care: Provider-reported rate and confidence. Fam Med 2007;39(10):730–5.

14. Leiserowitz A, Ballew M, Rosenthal S, et al. Climate change and the American diet. New Haven (CT): Yale Program on Climate Change Communication and Earth Day Network; 2020. Available: https://climatecommunication.yale.edu/wp-content/uploads/2020/02/climate-change-american-diet.pdf [Accessed 15 Jan 2021].

15. Neff RA, Edwards D, Palmer A, et al. Reducing meat consumption in the USA: A nationally representative survey of attitudes and behaviours. Public Health Nutr 2018;21(10):1835– 44.doi:10.1017/S1368980017004190

16. Dietary Guidelines Advisory Committee. Scientific Report of the 2020 Dietary Guidelines Advisory Committee: Advisory Report to the Secretary of Agriculture and the Secretary of Health and Human Services. Washington, DC: U.S. Department of Agriculture, Agricultural Research Service ; 2020. Available: https://www.dietaryguidelines.gov/2020-advisory-committee-report. [Accessed 20 Jan 2021].

17. Willett W, Rockström J, Loken B, et al. Food in the Anthropocene: the EAT–Lancet Commission on healthy diets from sustainable food systems. Lancet 2019;6736(18):3–49.doi:10.1016/S0140-6736(18)31788-4

18. Hemler EC, Hu FB. Plant-based diets for personal, population, and planetary health. Adv Nutr 2019;10(Suppl 4):PS275–83.doi:10.1093/advances/nmy117

19. Magkos F, Tetens I, Bügel SG, et al. A perspective on the transition to plant-based diets: A diet change may attenuate climate change, but can it also attenuate obesity and chronic disease risk? Adv Nutr 2020;11(1):1–9.doi:10.1093/advances/nmz090

20. Satija A, Hu FB. Plant-based diets and cardiovascular health. Trends Cardiovasc Med 2018;28(7):437–41.doi:10.1016/j.tcm.2018.02.004

21. Good Food Institute. U.S. retail market data for the plant-based industry, 2020. Available: https://www.gfi.org/marketresearch [Accessed 4 May 2021].

22. Mäkinen OE, Wanhalinna V, Zannini E, et al. Foods for special dietary needs: Non-dairy plant-based milk substitutes and fermented dairy-type products. Crit Rev Food Sci Nutr 2016;56(3):339–49.doi:10.1080/10408398.2012.761950

23. Vanga SK, Raghavan V. How well do plant based alternatives fare nutritionally compared to cow’s milk? J Food Sci Technol 2018;55(1):10–20.doi:10.1007/s13197-017-2915-y

24. Thorning TK, Raben A, Tholstrup T, et al. Milk and dairy products: Good or bad for human health? An assessment of the totality of scientific evidence. Food Nutr Res 2016;60:32527.doi:10.3402/fnr.v60.32527

25. Schuster MJ, Wang X, Hawkins T, et al. Comparison of the nutrient content of cow’s milk and nondairy milk alternatives: What’s the difference? Nutr Today 2018;53(4):153– 9.doi:10.1097/NT.0000000000000284

26. Chalupa-Krebzdak S, Long CJ, Bohrer BM. Nutrient density and nutritional value of milk and plant-based milk alternatives. Int Dairy J 2018;87:84–92.doi:10.1016/j.idairyj.2018.07.018

27. Ball L, Johnson C, Desbrow B, et al. General practitioners can offer effective nutrition care to patients with lifestyle-related chronic disease. J Prim Health Care 2016;5(1):59– 69.doi:10.1071/hc13059

28. Berhaupt A. The perceptions, attitudes and practices of registered dietitians regarding functional foods [master’s thesis]. Miami (FL): Florida International University; 2010.

29. Couch LM, Harris JE. Knowledge, attitudes, and self-reported practices of Pennsylvania registered dietitians regarding functional foods and herbal medicine. Top Clin Nutr 2008;23(1):32– 46.doi:10.1097/01.TIN.0000312078.45953.88

30. National Cancer Institute. Dietary Screener Questionnaires (DSQ) in NHANES 2009-10. Available: https://epi.grants.cancer.gov/nhanes/dietscreen/questionnaires.html [Accessed 31 Jan 2020].

31. Gresser MR. The consumption of dairy and dairy alternatives and the perception of dairy in college students [master’s thesis]. Kent (OH): Kent State University College; 2015.

32. Clarys P, Deliens T, Huybrechts I, et al. Comparison of nutritional quality of the vegan, vegetarian, semi-vegetarian, pesco-vegetarian and omnivorous diet. Nutrients 2014;6(3):1318– 32.doi:10.3390/nu6031318

33. USDA National Agriculture Statistics Service. Milk sales measured in % of farm sales, 2017. Availabile: https://quickstats.nass.usda.gov/results/AFA741E6-AAFC-302C-AB0F-08C82D5070E3/. [Accessed 28 April 2021].

34. Clark BE, Belarmino EH, Pope, L. Perspectives from healthcare professionals on the nutritional adequacy of plant-based dairy alternatives: Results of a mixed methods inquiry. Research Square [Preprint]. 2021 [cited 20 July 2021] doi:10.21203/rs.3.rs-726377/v1

35. U.S. Department of Health and Human Services and U.S. Department of Agriculture. Dietary Guidelines for Americans, 2020-2025. Available: https://www.dietaryguidelines.gov/ [Accessed 11 Jan 2021].

36. Ettinger J. 12,000 Doctors Urge the USDA to Issue Dairy Warning. LIVEKINDLY Collective, 2019. Available: https://www.livekindly.co/pcrm-dairy-dietary-guidelines-usda/ [Accessed 19 Nov 2020].

37. The Dairy Alliance. Health Benefits of Dairy. Available: https://thedairyalliance.com/dairy-nutrition/health-benefits-of-dairy/ [Accessed 31 May 2021].

38. Wolf CA, Malone T, McFadden BR. Beverage milk consumption patterns in the United States: Who is substituting from dairy to plant-based beverages? J Dairy Sci 2020;103(12):11209– 17.doi:10.3168/jds.2020-18741

39. Hess JM, Cifelli CJ, Fulgoni VL. Energy and nutrient intake of Americans according to meeting current dairy recommendations. Nutrients 2020;12(10):3006.doi: 10.3390/nu12103006

40. Bourassa L. Vegan and Plant-Based Diet Statistics for 2021. PlantProteins.co, 2021. Available: https://plantproteins.co/vegan-plant-based-diet-statistics/ [Accessed 6 May 2021].

41. Ipsos Retail Performance. Exploring the explosion of veganism in the United States. Available: https://www.ipsos-retailperformance.com/en/vegan-trends/ [Accessed 3 Aug 2020].

42. Aggarwal M, Singh Ospina N, Kazory A, et al. The mismatch of nutrition and lifestyle beliefs and actions among physicians: A wake-up call. Am J Lifestyle Med 2020;14(3):304– 15.doi:10.1177/1559827619883603

43. Priano SM, Hong OS, Chen JL. Lifestyles and health-related outcomes of U.S. hospital nurses: A systematic review. Nurs Outlook 2018;66(1):66–76.doi:10.1016/j.outlook.2017.08.013

44. Chen S, Kuhn M, Prettner K, et al. The macroeconomic burden of noncommunicable diseases in the United States: Estimates and projections. PLoS One 2018;13(11):e0206702.doi:10.1371/journal.pone.0206702

45. Food Insight. Survey: Nutrition Information Abounds, But Many Doubt Food Choices, 2017. Available: https://foodinsight.org/survey-nutrition-information-abounds-but-many-doubt-foodchoices/ [Accessed 20 May 2021].

46. Vinnari M, Montonen J, Härkänen T, et al. Identifying vegetarians and their food consumption according to self-identification and operationalized definition in Finland. Public Health Nutr 2009;12(4):481–8.doi:10.1017/S1368980008002486

47. Hebert JR, Clemow L, Pbert L, et al. Social desirability bias in dietary self-report may compromise the validity of dietary intake measures. Int J Epidemiol 1995;24(2):389– 98.doi:10.1093/ije/24.2.389

48. Commission on Dietetic Registration. Registry Statistics. Academy of Nutrition and Dietetics, 2021. Available: https://www.cdrnet.org/registry-statistics-new?id=1779&actionxm=ByDemographics [Accessed 24 May 2021].

49. Smiley RA, Lauer P, Bienemy C, Berg JG, Shireman E, Reneau KA, et al. The 2017 national nursing workforce survey. J Nurs Regul 2018;9(Suppl 3):PS1–88.doi:10.1016/S2155-8256(18)30131-5

50. U.S. Census Bureau. U.S. Census Bureau QuickFacts: Vermont, 2019. Available: https://www.census.gov/quickfacts/VT [Accessed 24 May 2021].

